# Serologic Responses to COVID-19 Vaccination in Children with History of Multisystem Inflammatory Syndrome (MIS-C)

**DOI:** 10.1101/2022.11.19.22282551

**Authors:** Maria A. Perez, Hui-Mien Hsiao, Xuemin Chen, Amber Kunkel, Nadine Baida, Laila Hussaini, Austin T. Lu, Carol M. Kao, Federico R. Laham, David A. Hunstad, Yajira Beltran, Teresa A. Hammett, Shana Godfred-Cato, Ann Chahroudi, Evan J. Anderson, Ermias Belay, Christina A. Rostad

## Abstract

Understanding the serological responses to COVID-19 vaccination in children with history of MIS-C could inform vaccination recommendations. We prospectively enrolled five children hospitalized with MIS-C and measured SARS-CoV-2 binding IgG antibodies to spike protein variants longitudinally pre- and post-Pfizer-BioNTech BNT162b2 primary series COVID-19 vaccination. We found that SARS-CoV-2 variant cross-reactive IgG antibodies waned following acute MIS-C, but were significantly boosted with vaccination and maintained for at least 3 months. We then compared post-vaccination binding, pseudovirus neutralizing, and functional antibody-dependent cell-mediated cytotoxicity (ADCC) titers to the reference strain (Wuhan-hu-1) and Omicron variant (B.1.1.529) among previously healthy children (n=6) and children with history of MIS-C (n=5) or COVID-19 (n=5). Despite the breadth of binding antibodies elicited by vaccination in all three groups, pseudovirus neutralizing and ADCC titers were reduced to the Omicron variant. Vaccination after MIS-C or COVID-19 (hybrid immunity) conferred advantage in generating pseudovirus neutralizing and functional ADCC antibodies to Omicron.

## INTRODUCTION

Multisystem inflammatory syndrome in children (MIS-C) associated with COVID-19 is a severe systemic inflammatory syndrome characterized by multiorgan involvement, myocardial dysfunction, shock, and occasionally death.^1^ The pathophysiology of MIS-C is poorly understood, but it is thought to represent a dysregulated immune response following infection with severe acute respiratory syndrome coronavirus-2 (SARS-CoV-2).^2,3^ Early studies found that MIS-C was associated with a robust serologic response to SARS-CoV-2.^4^ Nevertheless, significant declines in antibody titers are observed by 3 months,^5^ and the risk of reinfection with SARS-CoV-2 or recurrence of MIS-C following the initial diagnosis are unknown. Current Centers for Disease Control and Prevention (CDC) guidelines indicate that children with a history of MIS-C may receive COVID-19 vaccination, but should wait until at least 90 days after onset of MIS-C symptoms.^6^ In this study, we aimed to characterize the binding and functional serologic responses to COVID-19 vaccination in children with history of MIS-C.

## METHODS

### Patient Cohort

Following informed consent and assent, we prospectively enrolled hospitalized children who met the CDC case definition for MIS-C; children with PCR-confirmed symptomatic COVID-19; and healthy pediatric controls into a specimen collection protocol approved by the Institutional Review Board (IRB) at Emory University. Residual plasma and serum specimens were collected, and if the participant agreed, prospective blood was collected and the plasma and serum isolated. Hospitalized participants were invited to follow up longitudinally for repeat blood collection at 1, 3, 6, and 12 months post-hospitalization; and all participants were invited to follow up pre- and post-COVID-19 vaccination. Participants were included in this study if they had at least one specimen available at 1 month post-dose 2 of a primary BNT162b2 vaccine series.

### Serologic Analyses

Binding IgG antibodies to SARS-CoV-2 full-length spike from multiple strains, including wild-type (Wuhan-hu-1), Alpha (B.1.1.7), Beta (B.1.351), Delta (B.1.617.2), Gamma (P.1), and Omicron (B.1.1.529; BA.1), were measured using a MesoScale Discovery (MSD) V-PLEX SARS-CoV-2 panel per the manufacturer’s protocol. Titers were expressed as MSD arbitrary units (AU)/mL for spike variants in Fig. 1A-E and as WHO/NIBSC international standard binding antibody units (BAU)/mL for Wuhan-hu-1 spike in Fig. 1F. Binding IgG antibodies to the wild-type (Wuhan-hu-1) receptor binding domain (RBD) and nucleocapsid (N) protein were also measured using enzyme-linked immunosorbent assays (ELISAs) as previously described and expressed as end-point titers.^7^

**Figure 1.**
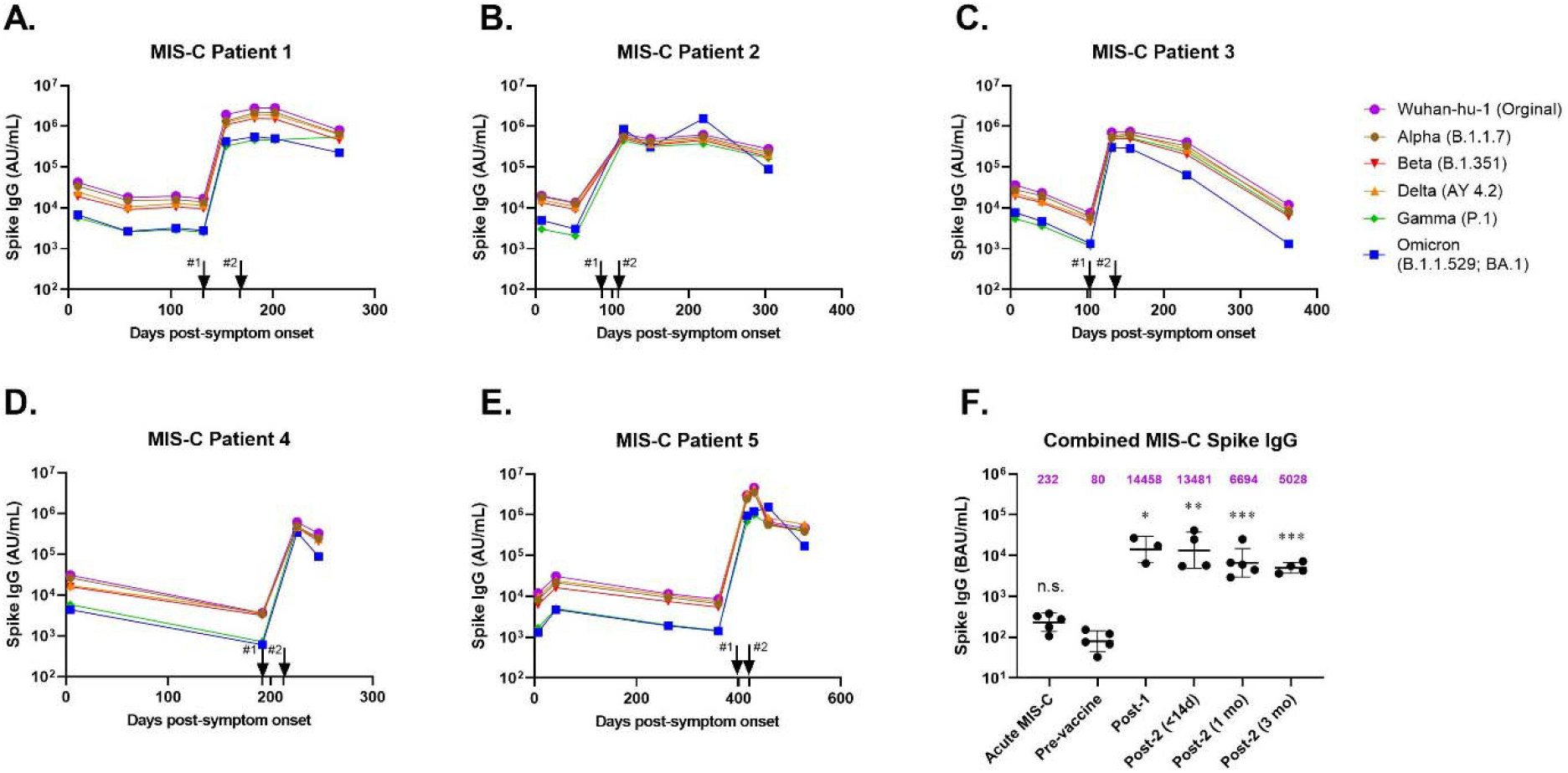
Longitudinal SARS-CoV-2 spike IgG antibodies in children with history of MIS-C following BNT162b2 primary series vaccination. Antibody titers are expressed in Mesoscale Discovery (MSD) arbitrary units (AU)/mL. Arrows represent vaccine doses #1 and #2. (F) SARS-CoV-2 spike (Wuhan-hu-1 strain) IgG in children with history of MIS-C during acute presentation with MIS-C, pre-vaccine, post-dose #1, post-dose #2 (<14 days), post-dose #2 (1 month), and post-dose #2 (3 months) with antibody titers expressed in WHO/NIBSC binding antibody units (BAU)/mL. Statistical comparisons (F) of log-transformed titers were performed using one-way ANOVA with Tukey’s post-hoc comparisons test comparing to pre-vaccine titers. Each dot represents an individual patient, and geometric means and geometric standard deviations are shown. \**P*≤ 0.05, \*\**P*≤ 0.01, \*\*\**P*≤ 0.005.

To perform pseudovirus neutralizing antibody assays, we developed lentiviral particles pseudotyped with either the wild-type (Wuhan-hu-1) or Omicron (B.1.1.529; BA.1) spike proteins.^8^ We incubated viral particles with serially diluted patient sera and applied the serum-virus mixture to 293T-ACE2 cells for 48 hours at 37°C. Following incubation, we added Britelite Plus (PerkinElmer) luciferase substrate and measured relative luminescence units (RLUs) to determine the effective concentration at which 50% of the virus was neutralized (EC50). The lower limit of detection (LLOD) was defined as the starting dilution of 1:30.

To perform antibody-dependent cell-mediated cytotoxicity (ADCC) assays as previously described,^8,9^ we generated stably transfected dual-reporter SARS-CoV-2 spike protein target cell lines with inducible expression of either wild-type (Wuhan-hu-1) or Omicron (B.1.1.529) spike proteins regulated by tetR in T-REx-293 cells. We then incubated the induced target cells with serially diluted patient sera for 1 hour at 37°C prior to adding NK-CD16^(+)^ effector cells and incubating for an additional 4 hours at 37°C. Following incubation, we added luciferase substrate and measured RLUs to determine the percent lysis of target cells. ADCC was calculated for each well using the following formula: ADCC (%) = [RLU (no serum) – RLU (with serum)]/RLU (no serum) x 100%. The serum dilution at which ADCC crossed the 10% cutoff was considered the end-point titer. The LLOD was defined as the starting dilution of 1:30.

### Statistical Analyses

Statistical analyses were performed in GraphPad Prism, v9.1.0. Baseline demographic characteristics were summarized using descriptive statistics. Geometric mean titers and geometric mean standard deviations were determined. All undetectable titers were assigned a value of ½ LLOD. Statistical comparisons of log-transformed titers were made using one-way analysis of variance (ANOVA) with Tukey’s post-hoc comparisons test. P-values <0.05 were considered statistically significant.

## RESULTS

Five children with MIS-C (three females, median age 15 years [IQR 13-16 years], date of symptom onset [DOS] range July 31, 2020 to February 24, 2021); five with COVID-19 (1 female, median age 15 years [IQR 14-16], DOS range November 29, 2020 to January 29, 2021); and 6 healthy pediatric controls (5 females, median age 15 years [IQR 13-15 years]) were enrolled and had post-vaccine samples available (Table 1). All children with MIS-C were hospitalized at the time of enrollment, 3 (60%) required intensive care, and the median duration of hospitalization was 5 days (IQR 5-7 days). Two children with COVID-19 (40%) were hospitalized, both required intensive care, and the median duration of hospitalization was 9 days (IQR 6-11 days). The timing of enrollment coincided with predominance of the D614G variant, and largely preceded circulation of the Alpha, Delta, and Omicron variants in the U.S.^10^ All children with MIS-C, two children with COVID-19, and one healthy control participant had samples available from multiple time points to allow for longitudinal analysis.

**Table 1.** Baseline demographic characteristics of cohort.

| **Variables** | **MIS-C** | **COVID-19** | **Healthy Controls** |
| --- | --- | --- | --- |
| **Number (n)** | 5 | 5 | 6 |
| **Female, n (%)** | 3 (60%) | 1 (20%) | 5 (83%) |
| **Age, median (IQR)** | 15 (13-16) | 15 (14-16) | 15 (13-15) |
| **Race, n (%)** |  |  |  |
| **Asian** | 1 (20%) | 0 (0%) | 1 (17%) |
| **Black** | 1 (20%) | 1 (20%) | 1 (17%) |
| **White** | 2 (40%) | 2 (40%) | 2 (33%) |
| **Multiple race** | 0 (0%) | 0 (0%) | 2 (33%) |
| **Declined** | 1 (20%) | 2 (40%) | 0 (0%) |
| **Ethnicity, n (%)** |  |  |  |
| **Hispanic** | 2 (40%) | 4 (80%) | 1 (17%) |
| **Hospitalized** | 5 (100%) | 2 (40%) | --- |
| **ICU** | 3 (60%) | 2 (40%) | --- |
| **Duration of hospitalization, median (IQR)** | 5 (5-7) | 9 (6-11) | --- |
| **Time from symptom onset to vaccination, median (IQR)** | 132 (104-192) | 143 (130-171) | --- |
| **Dates of symptom onset (range)** | 7/31/2020-2/24/2021 | 11/29/2020-1/29/2021 | --- |
| **Dates of vaccine dose#1 (range)** | 4/9/2021-12/7/2021 | 5/19/2021-7/18/2021 | 5/12/2021-6/1/2021 |
| **Dates of vaccine dose#2 (range)** | 4/30/2021-12/26/2021 | 6/9/2021-8/9/2021 | 6/2/2021- 6/16/2021 |

Binding IgG antibodies to SARS-CoV-2 full-length spike variants were elevated during the acute hospitalization for MIS-C (Figure 1) and COVID-19 (Supplementary Figure 1), but waned prior to vaccination,^11^ especially for Gamma and Omicron (Figure 1). Following vaccination, broadly cross-reactive binding IgG antibodies were boosted and maintained for up to 3 months. For the wild-type (Wuhan-hu-1) spike, at a median follow-up of 132 days (IQR 104-192 days) from MIS-C symptom onset, pre-vaccine binding IgG GMT was 80 BAU/mL (95% confidence interval [CI] 38 to 167) (Figure 1F). IgG titers increased significantly following 1 dose (GMT 14458, 95%CI 2333 to 89616; P=0.019) and 2 doses (GMT 13481, 95%CI 2633 to 69029; P=0.008) of BNT162b2 vaccine. These titers were maintained at 1 month (GMT 6694, 95%CI 2457 to 18237; P<0.005) and 3 months (GMT 5028, 95%CI 3113 to 8120; P<0.005) post-vaccination. Similar trends were observed with binding IgG antibodies to the spike RBD as measured by ELISA, as vaccination elicited significant increases in titers in all 5 children with history of MIS-C (Supplementary Figure 2).

We then compared the post-vaccination binding, pseudovirus neutralizing, and ADCC titers to wild-type vs. Omicron variant in children with history of MIS-C, COVID-19, and healthy pediatric controls. Samples were collected approximately 1 month following dose 2 of the BNT162b2 primary series for children with history of MIS-C (median 34 days; IQR 33-38 days); COVID-19 (median 40 days; IQR 30-42 days); and healthy pediatric controls (median 34 days; IQR 33-35 days). Post-vaccination binding IgG titers to wild-type (Wuhan-hu-1) spike were similar between the cohorts (Figure 2A). Binding antibodies to the Omicron spike were reduced 2-to 4-fold compared to wild-type spike for all groups, but these differences were not statistically significant.

**Figure 2.**
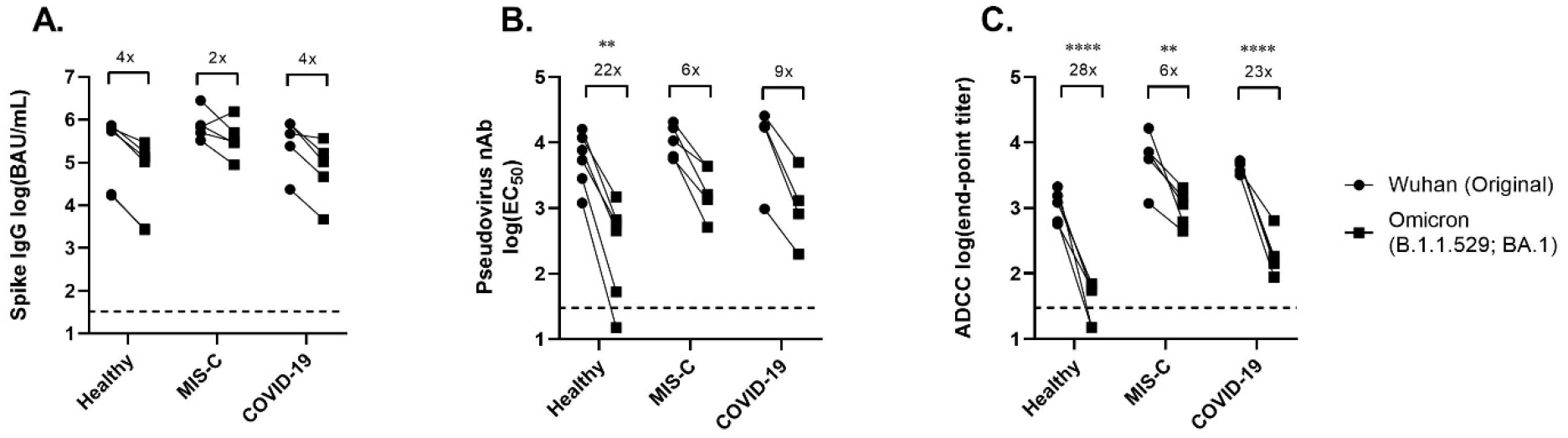
SARS-CoV-2 antibody responses to Wuhan vs. Omicron (BA.1) variant in children post-BNTb162b vaccine primary series. (A) Spike IgG (AU/mL); (B) Pseudovirus neutralizing antibodies (EC50); and (C) Antibody-dependent cell-mediated cytotoxicity (ADCC) antibody end-point titers are shown for children who were previously healthy, children with history of MIS-C, and children with history of COVID-19. The dashed line represents the assay lower limit of detection (LLOD). Fold-change in the geometric mean titers of Wuhan vs. Omicron strains are shown. Statistical comparisons of log-transformed titers were performed using two-way ANOVA with Tukey’s post-hoc comparisons test. AU, arbitrary units. EC50, estimated serum dilution at which 50% of pseudovirus is neutralized. \*\**P*≤ 0.01, \*\*\*\**P*≤ 0.001.

Post-vaccination pseudovirus neutralizing antibody titers to the wild-type strain were also similar between the cohorts (Figure 2B). However, pseudovirus neutralizing titers to the Omicron variant were significantly reduced in the healthy control group (22-fold; Wuhan-hu-1 GMT 5472, 95%CI 1997 to 14989 vs. Omicron GMT 243, 95% CI 38 to 1551; P=0.0027). Pseudovirus neutralizing titers to Omicron were more modestly reduced in the children with history of MIS-C (6-fold; Wuhan-hu-1 GMT 10525, 95%CI 5126 to 21610 vs. Omicron GMT 1826, 95%CI 604 to 5511; P=0.260) and COVID-19 (9-fold; Wuhan-hu-1 GMT 9371, 95%CI 834 to 105259; vs. Omicron GMT 1014, 95%CI 122 to 8406; P=0.153), but these differences were not statistically significant.

Post-vaccination functional ADCC titers to the Wuhan-hu-1 strain were significantly higher in children with history of MIS-C (GMT 5597, 95%CI 1692 to 18519) or COVID-19 (GMT 4318, 95%CI 3296 to 5658) when compared to healthy pediatric controls (GMT 1100, 95%CI 639 to 1893; P=0.006 and P=0.027, respectively). ADCC titers to the Omicron variant were significantly reduced in all three groups, including the healthy pediatric controls (28-fold; GMT 39, 95%CI 18-86; P<0.001); children with history of MIS-C (6-fold; GMT 986, 95%CI 453 to 2145; P=0.005); and children with history of COVID-19 (23-fold; GMT 187, 95%CI 75 to 471, P<0.001).

## DISCUSSION

We analyzed the serologic responses to SARS-CoV-2 wild-type strain and variants among children with a history of MIS-C, COVID-19, and healthy pediatric controls who received a BNT162b2 vaccine primary series. Children with MIS-C and COVID-19 had elevated SARS-CoV-2 spike IgG antibodies against Wuhan-hu-1 during acute hospitalization that waned prior to vaccination. Administration of a BNT162b2 primary series boosted broadly cross-reactive binding antibodies in these children which were maintained for at least 3 months. Despite improvements in the breadth of binding antibodies elicited by vaccination in all three groups, pseudovirus neutralizing and ADCC titers tended to be reduced against the Omicron variant. Prior infection, either resulting in COVID-19 or MIS-C, followed by vaccination (e.g., hybrid immunity) resulted in broader pseudovirus neutralizing and functional ADCC titers to Omicron than did the 2-dose primary vaccination series alone.

Currently, the CDC advises that children with a history of MIS-C may receive COVID-19 vaccination, but that they should wait at least 90 days following resolution of MIS-C symptoms.^6^ The rationale for this delayed interval has been that children with MIS-C are likely to have increasing risk of re-infection (with the possibility of recurrent MIS-C) as more time elapses from their MIS-C hospitalization. This risk has been balanced with the unknown risks of adverse effects to vaccination in this unique population. While MIS has been reported in children^12^ and adults^13,14^ following COVID-19 vaccination, this has been an exceedingly rare occurrence. Further, a recent multicenter U.S. study demonstrated that a 2-dose primary BNT162b2 vaccine series was 91% effective (95%CI 78% to 97%) in preventing MIS-C.^15^ Importantly, no children with history of MIS-C in our study developed recurrent MIS-C or any adverse events requiring medical attention following COVID-19 vaccination.

Recent studies have illuminated immune correlates of protection from COVID-19 illness and hospitalization. For example, an analysis of the ChAdOx1 nCoV-19 (AZD1222) vaccine found that a spike IgG titer of 264 BAU/mL (95%CI 108-806) correlated with a vaccine efficacy of 80% against symptomatic infection with majority Alpha (B.1.1.7) variant.^11^ Other correlates of protection identified in that study included RBD binding antibodies, pseudovirus neutralizing, and live-virus neutralizing antibodies. An analysis of the mRNA-1273 phase 3 efficacy trial found similar correlates of protection and estimated that neutralizing antibodies mediate approximately two-thirds of mRNA-1273 vaccine efficacy.^16^ Goldblatt, et. al. utilized population-based methods to assess the correlation of IgG binding with vaccine efficacy to estimate a protective threshold against SARS-CoV-2 wild-type (154 BAU/mL, 95%CI 42 to 559) and Alpha (B.1.1.7) (171 BAU/mL, 95%CI 57 to 519).^17^ Nevertheless, a correlate of protection against the Omicron variant has not been established, and the contributions of other humoral responses, such as functional ADCC antibodies, memory B cells, and T cell responses to protective immunity are incompletely understood.

ADCC is one of several functional, non-neutralizing antibody responses elicited by SARS-CoV-2 infection.^9,18,19^ While functional antibodies have been found to correlate with COVID-19 severity^18^ and mortality^19^ in adults, little is known about their role in protection against disease in children. Children with hybrid immunity (prior infection plus vaccination) in our study had significantly higher functional ADCC responses to both the wild-type (Wuhan-hu-1) strain and the Omicron variant, despite having non-significant differences in binding and pseudovirus neutralizing titers. As hybrid immunity has previously been reported to reduce the incidence SARS-CoV-2 re-infection and hospitalization,^20^ future studies are needed to elucidate the role of ADCC in protection.

Our study has some limitations. This was a single-center analysis with a small sample size, and longitudinal samples were lacking for several participants. The infecting strain was not known for children with previous COVID-19 or MIS-C. T and memory B cell responses, non-spike ADCC responses, and the effects of booster vaccination were not ascertained.

In conclusion, following hospitalization for MIS-C, binding IgG antibodies waned prior to administration of COVID-19 vaccine. A primary BNT162b2 series elicited broadly cross-reactive antibodies that persisted for at least 3 months. Hybrid immunity (prior infection plus vaccination) conferred an advantage in terms of functional ADCC antibody responses. Future studies are needed to describe the safety and reactogenicity of vaccination in this unique patient population and the correlates of protection.

## Data Availability

All data produced in the present study are available upon reasonable request to the authors.

## ACKNOWLEDGEMENTS

We thank Dr. Jens Wrammert for kindly sharing purified RBD protein antigen. We thank clinical research coordinators Felicia Glover, Vikash Patel, Amber Samuel, and clinical research nurses Lisa Macoy and Kathy Stephens for their assistance enrolling patients and collecting specimens. We thank Theda Gibson, Wensheng Li, and the Emory Children’s Center Vaccine Research Clinic (ECC-VRC) laboratory for their assistance processing specimens. We thank the Children’s Healthcare of Atlanta research laboratory for their assistance in collecting residual specimens. We also thank the study participants and their families for generously donating their time and blood to further our understanding of COVID-19 and MIS-C.

## DISCLAIMER

The findings and conclusions in this report are those of the authors and do not necessarily represent the official position of the Centers for Disease Control and Prevention.

## FUNDING STATEMENT

This work was funded by Emory University and Children’s Healthcare of Atlanta, and by the U.S. Centers for Disease Control and Prevention.

§ See e.g., 45 C.F.R. part 46, 21 C.F.R. part 56; 42 U.S.C. §241(d); 5 U.S.C. §552a; 44 U.S.C. §3501 et seq.

## PRIOR PRESENTATIONS

The information contained herein has not been previously published or presented at any meetings.

