## Supplementary data for "Serologic Responses to COVID-19 Vaccination in Children with History of Multisystem Inflammatory Syndrome (MIS-C)"

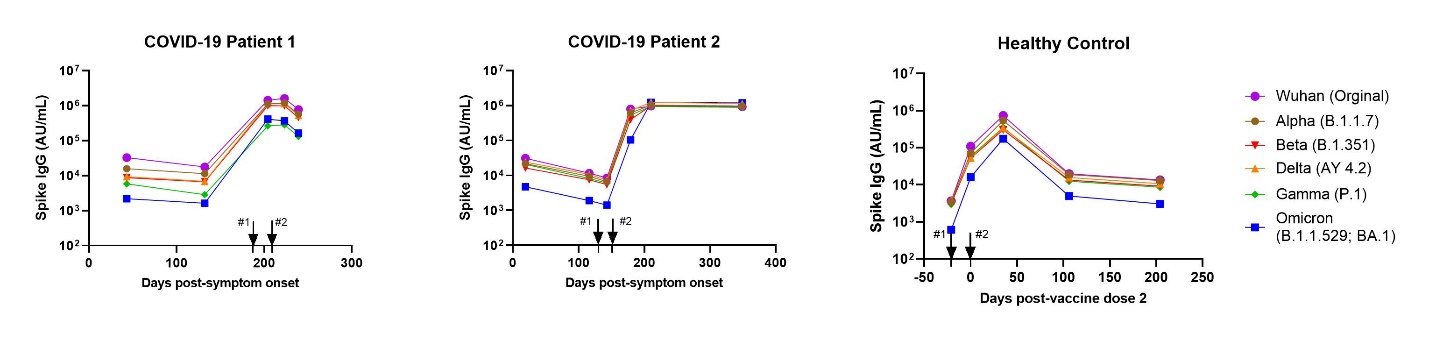

**Supplemental Figure 1.** **Longitudinal SARS-CoV-2 variant spike IgG antibodies in children with history of COVID-19 or a Healthy Control following BNT162b2 vaccination.** Arrows represent vaccine doses #1 and #2, respectively. Antibody titers are expressed in Mesoscale Discovery (MSD) arbitrary units (AU)/mL. The LLOD is <10^2^ AU/mL for all variants.

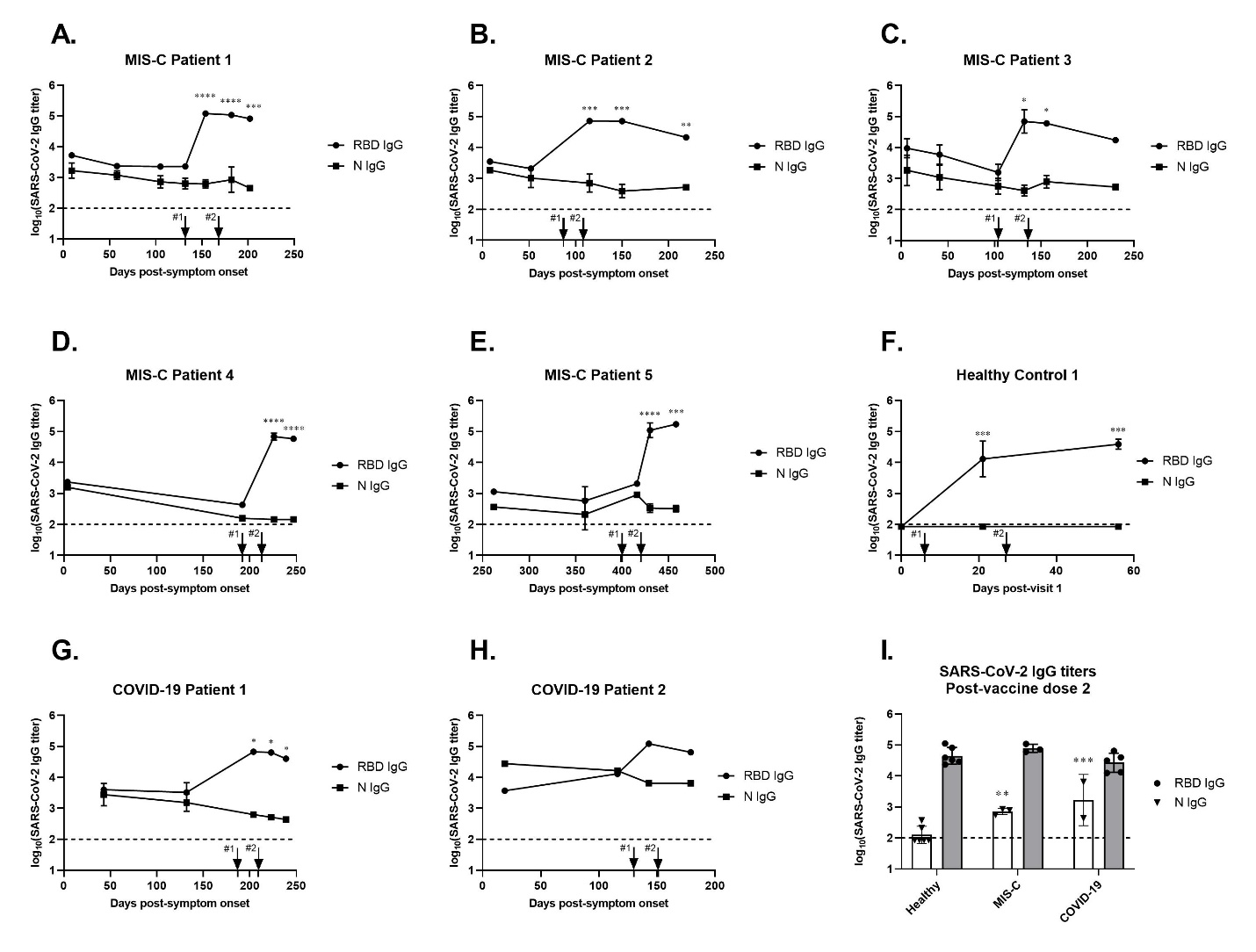

**Supplemental Figure 2.** **Longitudinal SARS-CoV-2 receptor binding domain (RBD) and nucleocapsid (N) protein IgG binding antibodies by ELISA in children with history of MIS-C, healthy control, or COVID-19 following BNT162b2 vaccination.** The x-axes represent the time post-onset of MIS-C symptoms (A-E), time post-sample 1 (F), or time post-onset of COVID-19 symptoms (G-H). Arrows represent vaccine doses #1 and #2, respectively. (I) Post-vaccine SARS-CoV-2 RBD and N IgG end-point titers for children who were previously healthy and children with history of MIS-C or COVID-19. Statistical comparisons of log-transformed titers were performed using one-way ANOVA with Tukey’s post-hoc comparisons test comparing to healthy controls. Each dot represents an individual patient, and geometric means and geometric standard deviations are shown. ***P*≤0.01, ****P*≤0.005.
